# Childhood Maltreatment and Mental-Physical Multimorbidity: An Analysis of Canadian Survey Data Using Entropy Balancing

**DOI:** 10.1101/2025.05.15.25326936

**Authors:** Andrew J. Fullerton, Alpamys Issanov, Rafael Meza

## Abstract

**Objectives:** To explore childhood maltreatment as a risk factor for mental-physical multimorbidity and examine gender as an effect modifier.

**Methods:** We analyzed data from the 2022 Mental Health and Access to Care Survey. We described sample characteristics with unweighted counts, survey-weighted percentages, and weighted chi-square tests. Missing data were addressed via multiple imputation. Entropy balancing adjusted for age, gender, LGBTQ2+ identity, visible minority group, and immigration status and multinomial logistic regression was used to estimate associations between the number of childhood maltreatment subtypes (physical abuse, sexual abuse, and exposure to domestic violence) reported and physical (≥ 2 physical conditions but no mental), mental (≥ 2 mental conditions but no physical), and mental-physical (≥ 1 mental and physical condition) multimorbidity. Survey weights were applied during both entropy balancing and regression modeling. Effect modification by gender was examined and sub-analyses of mental- cardiometabolic, mental-inflammatory, mental-somatic multimorbidity, and subtype-specific exposures were conducted.

**Results:** 8,967 respondents were included. Mental-physical multimorbidity increased with maltreatment: 3.4% (none, n=4647), 6.3% (1 type, n=2804), 10.1% (2 types, n=1208), and 18.2% (3 types, n=308). Adjusted odds ratios for mental-physical multimorbidity ranged from 2.15 (95% CI:1.90-2.44) for 1 type to 8.72 (95% CI:7.01-10.85) for 3 types compared to physical (aOR=1.31-2.00) and mental (aOR=1.90-3.63) multimorbidity. Men showed higher odds of mental-physical multimorbidity at high exposure (aOR=6.14, 95% CI:4.90-7.70 in women; aOR=13.96, 95% CI:9.58-20.34 in men) with varying effect sizes across disease areas.

**Conclusion:** Childhood maltreatment shows a strong dose-response association with mental-physical multimorbidity. Further research is needed to clarify gender-specific pathways.

## INTRODUCTION

Multimorbidity has been shown to worsen the prognosis of all present diseases while making treatment more complicated, more costly, and less effective. These challenges are even greater for individuals with co-occurring mental and physical health conditions due to limited clinical overlap between non-psychiatric and psychiatric specialists leading to fragmented care that does not address the interacting effects of both conditions (Halstead et al., 2024). With an estimated prevalence of 8.4% in Canada as of 2014, individuals with mental-physical multimorbidity – the co-occurrence of at least one mental and one physical chronic health condition – report higher rates of suicidal ideation, more adverse health outcomes, greater healthcare utilization and more unmet healthcare needs across the life course compared to those with only physical or mental multimorbidity (Dai et al., 2020). Given its substantial burden, identifying modifiable early life risk factors for mental-physical multimorbidity is an important public health priority. While childhood maltreatment is an established early life risk factor for various mental and physical health conditions, its role in mental-physical multimorbidity remains underexplored in Canada.

Childhood maltreatment is thought to affect a variety of health outcomes through increased risk of health-harming behaviours and chronic dysregulation of neuroregulatory pathways such as the hypothalamic-pituitary-adrenal (HPA) axis (Hughes et al., 2017; Metzler et al., 2016). Recent studies using data from the Canadian Community Health Survey (CCHS) have validated the association between childhood maltreatment and increased risk of diabetes (Shields et al., 2016a), chronic obstructive pulmonary disease (Shields et al., 2016b), arthritis (Badley et al., 2019), cancer (Hovdestad et al., 2020), and mental health conditions including anxiety, depression, and substance use disorder (Afifi et al., 2014). Other research in Canada has identified childhood maltreatment as a key risk factor for psychiatric comorbidity in individuals with multiple sclerosis, arthritis, and inflammatory bowel disease (Wan et al., 2022; O’Mahony et al., 2024). To our knowledge, however, only one study (England-Mason et al., 2018) has assessed childhood maltreatment as a risk factor for multimorbidity across health domains in a representative sample of the Canadian population. Consistent with other evidence, their findings suggest a dose-response relationship between the number of maltreatment types experienced and multimorbidity in adulthood.

While England-Mason et al. (2018) provided initial evidence of the association, the role of childhood maltreatment in mental-physical multimorbidity has not been explored in-depth. Despite recent evidence indicating climbing multimorbidity rates among Canadian men (Kone et al., 2021; Ferris et al., 2025), no work has been published examining potential gender differences. Our study addresses this gap by examining the association between childhood maltreatment and mental-physical multimorbidity using weighted, nationally representative data from the 2022 Mental Health and Access to Care Survey (MHACS). This work aims to provide a foundation for future research into the early life mechanisms underlying mental-physical multimorbidity and inform more targeted prevention efforts. Building on England-Mason et al. (2018), we aim to explore childhood maltreatment as a risk factor for mental-physical multimorbidity across disease areas and examine gender as a potential effect modifier.

## METHODS

### Data Source

MHACS is a multi-stage stratified survey designed to assess the mental health, service use, and functioning of Canadians including individuals aged 15 and older as of March 1, 2022, residing in the ten Canadian provinces (Statistics Canada, 2023). The sampling frame is representative of the Canadian population excluding the Canadian territories, full-time Canadian Forces members, individuals living on reserves or Indigenous settlements, and those residing in collective dwellings. Out of 39,485 units selected from the 2021 census, 9,861 individuals responded to the survey, yielding an overall response rate of around 25%. Data were collected between March 2022 and July 2022 using computer-assisted telephone interviews. Further details are available from Statistics Canada. All data used in this analysis were obtained through the Abacus Data Network (Abacus Data Network, n.d.).

### Analytic Sample

The sample included respondents who were administered the childhood experiences section of MHACS. Invalid responses and non-responses were imputed using multiple imputation, resulting in an analytic sample of 8,967 individuals aged 20 and older.

### Study Variables

#### Childhood Maltreatment

For consistency with prior research and the Canadian Child Maltreatment Surveillance Indicator framework (Campeau et al., 2020), we derived an exposure variable measuring the number of childhood maltreatment subtypes reported ranging from none to three (physical abuse, sexual abuse, and exposure to domestic violence). Respondents reported childhood maltreatment experiences before age 16 using items from the Childhood Experiences of Violence Questionnaire (Walsh et al., 2008) with additional items for sexual abuse (Statistics Canada, 2023). Physical abuse was classified as present if respondents reported at least one occurrence of being physically attacked. Sexual abuse was classified as present if respondents reported at least one instance of forced unwanted sexual activity or touching. Exposure to domestic violence was classified as present if respondents reported witnessing a parent or guardian physically harm another adult in the home.

#### Mental-Physical Multimorbidity Status

We derived the outcome variable, mental-physical multimorbidity status, based on self-reported reported chronic conditions across two domains: mental and physical health. To enable comparisons between mental-physical and single-domain multimorbidity, the variable was constructed as a multilevel nominal variable with four states: no multimorbidity (reference group), physical multimorbidity (≥ 2 physical conditions but no mental), mental multimorbidity (≥ 2 mental conditions but no physical), and mental-physical multimorbidity (≥ 1 mental and physical condition).

Mental health conditions were assessed using the WHO 12-Month Composite International Diagnostic Interview criteria (Statistics Canada, 2023) and included major depression, generalized anxiety disorder, social phobia, bipolar I/II, hypomania, and substance use disorders (alcohol, cannabis, and other drugs). Physical health conditions were self-reported and included hypertension, diabetes, heart disease, asthma, arthritis, inflammatory bowel disease (IBD), chronic obstructive pulmonary disease (COPD), chronic fatigue syndrome, fibromyalgia, back pain, cancer, and chronic migraines.

#### Covariates

We used a directed acyclic graph (Fig 1) to represent hypothesized relationships between variables and identify a minimally sufficient adjustment set: age, gender, LGBTQ2+ identity, visible minority group, and immigration status. Health-related behaviours and adulthood socioeconomic factors were identified as mediators and omitted from the analytic model to allow total effect estimation.

**Fig 1.**
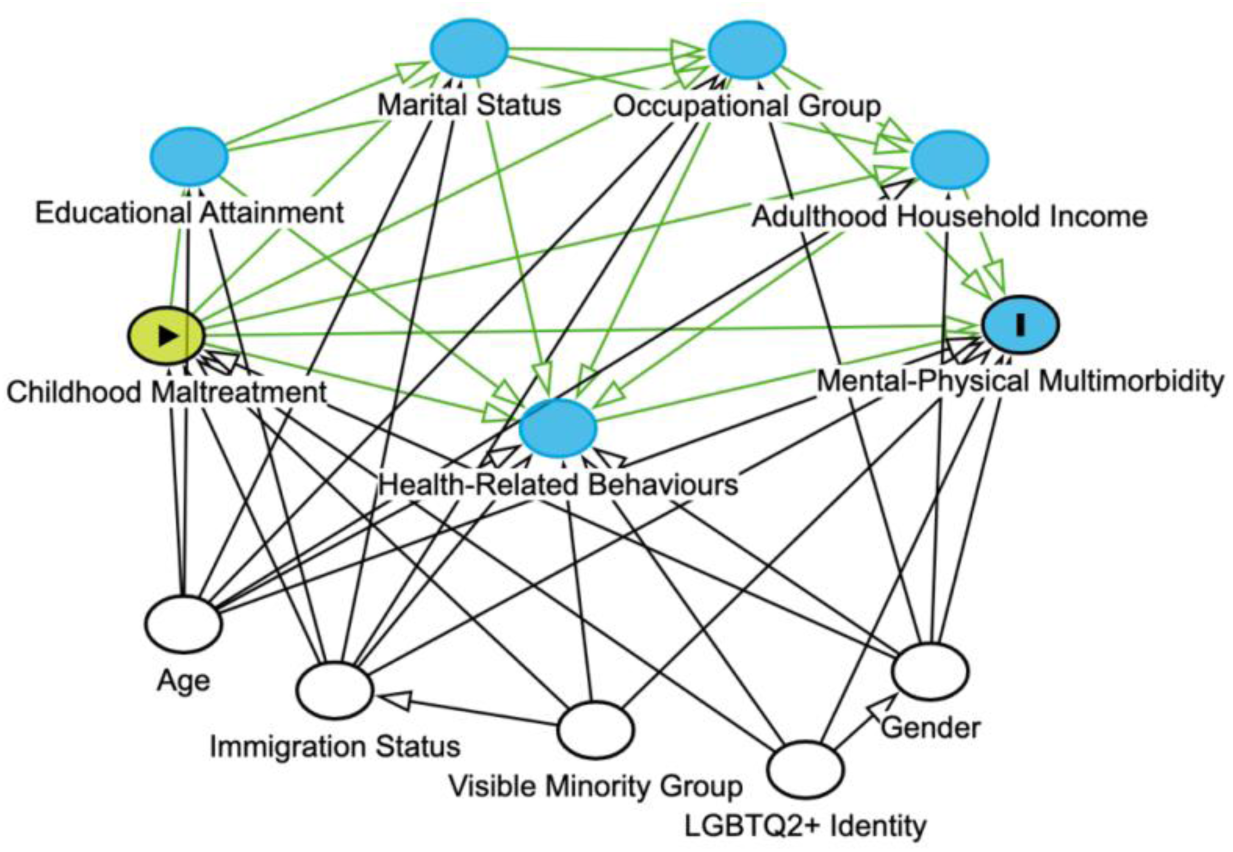
Directed acyclic graph presenting relationships between exposure, outcome, and measured covariates based on a priori understanding. Blue nodes indicate mediator variables while green edges indicate causal paths. White nodes indicate adjusted variables and black edges indicate blocked paths.

### Statistical Analysis

We described respondent characteristics and multimorbidity prevalence using unweighted counts and survey-weighted percentages, stratified by the number of childhood maltreatment subtypes reported. We assessed associations between characteristics and exposure groups using survey-weighted chi-square tests.

To adjust for confounding, we used entropy balancing to create a reweighted pseudopopulation balanced on age, gender, LGBTQ2+ identity, visible minority group, and immigration status.

Entropy balancing is a reweighting method that achieves covariate balance by directly matching covariate distributions across treatment groups, ensuring balance on specified moments (e.g., mean, variance, and skewness) while minimizing divergence from uniform base weights via an optimization function (Hainmueller, 2012). Since all included covariates were categorical, balance was achieved on proportions with a constraint tolerance of 1e-10 and 1000 maximum iterations.

We estimated associations between childhood maltreatment and mental-physical multimorbidity status using multinomial logistic regression. To examine effect modification, we added a gender-by-maltreatment interaction after confirming covariate balance across gender (standardized mean difference < 0.1). Sampling weights were applied during both entropy balancing and regression modeling to maintain population representativeness. Variance was estimated using a bootstrap estimator with replicate weights provided by Statistics Canada to account for survey design. All analyses were conducted in R 4.5.1.

### Missing Data

After determining that data were not missing completely at random through exploratory visualization, data were assumed to be missing at random in approximately 2.41% of the unweighted sample (Table 1). Missingness was modest (less than 5%) across all variables except for mental health conditions where missingness was 12%.

**Table 1.** Characteristics of Canadian adults (> 20 years of age) stratified by the number of types of childhood maltreatment experiences reported before 16 years of age based on the 2022 Mental Health and Access to Care Survey (MHACS).

| Variable | None<sup>1</sup><br>(n = 4647) | 1 maltreatment<br>type<sup>1</sup><br>(n = 2804) | 2 maltreatment<br>types<sup>1</sup><br>(n = 1208) | 3 maltreatment<br>types<sup>1</sup><br>(n = 308) | p-value |
| --- | --- | --- | --- | --- | --- |
| **Age (%)** |  |  |  |  | <0.001<sup>2</sup> |
| 20 to 24 years | 643 (9.8) | 307 (7.1) | 169 (7.8) | 34 (7.0) |  |
| 25 to 29 years | 291 (7.7) | 175 (6.0) | 69 (5.5) | 21 (7.3) |  |
| 30 to 34 years | 359 (9.0) | 181 (7.4) | 84 (8.0) | 23 (7.9) |  |
| 35 to 44 years | 678 (16.8) | 465 (19.1) | 190 (17.6) | 46 (15.5) |  |
| 45 to 54 years | 600 (13.6) | 366 (14.3) | 176 (15.3) | 61 (18.9) |  |
| 55 to 64 years | 647 (17.0) | 437 (19.6) | 196 (21.3) | 54 (23.5) |  |
| 65 years or older | 1,429 (26.0) | 873 (26.4) | 324 (24.5) | 69 (19.9) |  |
| **Gender (%)<sup>3 4</sup>** |  |  |  |  | <0.001<sup>2</sup> |
| Women+ | 2,439 (54.3) | 1,193 (44.4) | 575 (50.0) | 219 (73.6) |  |
| Men+ | 2,200 (45.7) | 1,607 (55.6) | 631 (50.0) | 89 (26.4) |  |
| **LGBTQ2+** |  |  |  |  | <0.001<sup>2</sup> |
| Cisgender/Heterosexual | 4,242 (95.8) | 2,624 (94.3) | 1,112 (92.5) | 257 (82.7) |  |
| LGBTQ2+ | 185 (4.2) | 152 (5.7) | 84 (7.5) | 47 (17.3) |  |
| **Visible minority group (%)<sup>3</sup>** |  |  |  |  | <0.001<sup>2</sup> |
| Not a visible minority | 2,382 (72.6) | 1,576 (76.5) | 607 (70.8) | 166 (72.4) |  |
| Black | 377 (3.1) | 305 (3.7) | 163 (5.3) | 34 (3.9) |  |
| Chinese | 446 (5.6) | 281 (5.3) | 107 (4.1) | 28 (5.2) |  |
| Filipino | 381 (2.8) | 177 (2.1) | 87 (2.9) | 19 (2.5) |  |
| South Asian | 500 (8.5) | 225 (5.7) | 136 (8.9) | 27 (6.5) |  |
| Other visible minority | 308 (7.4) | 183 (6.7) | 91 (8.1) | 28 (9.5) |  |
| **Immigration status (%)<sup>3</sup>** |  |  |  |  | 0.2<sup>2</sup> |
| Immigrant | 2,023 (32.1) | 1,187 (29.6) | 546 (32.4) | 124 (31.4) |  |
| Canadian-born | 2,419 (67.9) | 1,600 (70.4) | 658 (67.6) | 183 (68.6) |  |

Unweighted counts and survey-weighted percentages are reported; column totals reflect unweighted population including missingness.
<sup>1</sup> Maltreatment types included are physical abuse, sexual abuse, and exposure to domestic violence.
<sup>2</sup> Weighted chi-square test used to calculate p-values.
<sup>3</sup> Missing: Gender (0.15%); Visible minority group (3.75%); Immigration status (2.36%); LGBTQ2+ (2.99%).
<sup>4</sup> Mental Health and Access to Care Survey (MHACS) used random imputation to distribute non-binary persons into either Men+ or Women+.

We addressed missing values using multiple imputation by chained equations with 10 imputations based on convergence and 5 iterations to reduce error in the pooled estimates. The imputation model included all non-derived study variables and auxiliary variables (self-rated mental health, self-rated physical health, family member mental health, household income, occupational status, marital status, and educational attainment) to improve imputation of outcome variables and support the missing at random assumption. Following imputation, derived variables were created and entropy balancing and regression modeling were performed on each imputed dataset. Final estimates were pooled using Rubin’s rules (Rubin, 1987).

### Cluster-Specific Analyses

To examine variations across previously identified clusters of co-occurring mental (including substance use disorders) and physical disease, we conducted sub-analyses of mental- cardiometabolic (co-occurring mental illness and diabetes, heart disease, or hypertension), mental-inflammatory (co-occurring mental illness and asthma, arthritis, COPD, or IBD), and mental-somatic multimorbidity (co-occurring mental illness and chronic fatigue syndrome, back pain, chronic migraines, or fibromyalgia). Clusters were constructed based on shared risk factors, pathophysiology, and previously identified links to childhood maltreatment (Baltramonaityte et al., 2023; Souama et al., 2023; Wan et al., 2022; O’Mahony et al., 2024; England-Mason et al., 2018).

### Subtype-Specific Analyses

To investigate subtype-specific effects, maltreatment subtypes were coded as binary variables (reported/not reported) and separate models were created for each maltreatment subtype with additional adjustment for the non-focal subtypes.

### Ethics Approval

This secondary data analysis did not require institutional ethics review (Canadian Institutes of Health Research, 2022).

## RESULTS

### Descriptive Statistics

Out of 9,861 MHACS respondents, 8,967 individuals (90.9%) were included in the analysis. Age, visible minority group, and immigration status distributions were consistent across all levels of exposure to childhood maltreatment (Table 1). Notably, the proportion of women increased with exposure to childhood maltreatment (1 type: 44.4%, 2 types: 50.0%, 3 types: 73.6%) as did the proportion identifying as LGBTQ2+ (1 type: 5.7%, 2 types: 7.5%, 3 types: 17.3%).

Table 2 reports the estimated prevalence of mental-physical multimorbidity (5.7%), mental multimorbidity (10.7%), and physical multimorbidity (24.8%) in the Canadian population. The prevalence of mental-physical multimorbidity increased from 3.4% among those who reported no maltreatment (n=4647) to 18.2% among those who reported 3 maltreatment types (n=308), showing a clear gradient. Physical multimorbidity was elevated in those who reported any maltreatment (1 type: 26.8%; 2 types: 27.5%; 3 types: 25.8%) compared to none (22.7%). Mental multimorbidity prevalence increased steadily with exposure to childhood maltreatment from 8.2% (no maltreatment) to 18.6% (3 types).

**Table 2.**
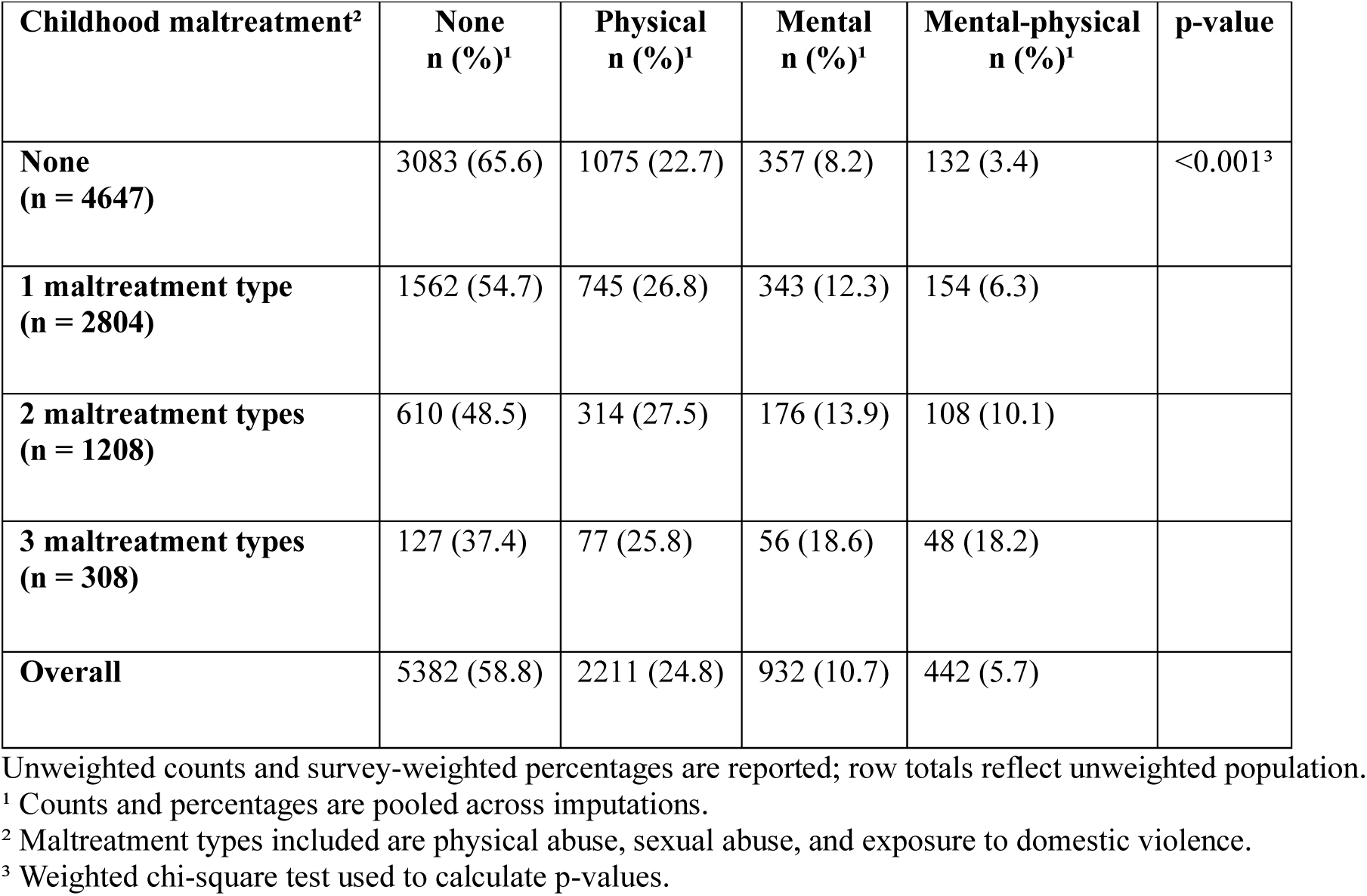
Prevalence of multimorbidity by number of childhood maltreatment types experienced in Canadian individuals 20 years of age and older based on the 2022 Mental Health and Access to Care Survey (MHACS).

| Childhood maltreatment<sup>2</sup> | None<br>n (%)<sup>1</sup> | Physical<br>n (%)<sup>1</sup> | Mental<br>n (%)<sup>1</sup> | Mental-physical<br>n (%)<sup>1</sup> | p-value |
| --- | --- | --- | --- | --- | --- |
| **None**<br>**(n = 4647)** | 3083 (65.6) | 1075 (22.7) | 357 (8.2) | 132 (3.4) | <0.001<sup>3</sup> |
| **1 maltreatment type**<br>**(n = 2804)** | 1562 (54.7) | 745 (26.8) | 343 (12.3) | 154 (6.3) |  |
| **2 maltreatment types**<br>**(n = 1208)** | 610 (48.5) | 314 (27.5) | 176 (13.9) | 108 (10.1) |  |
| **3 maltreatment types**<br>**(n = 308)** | 127 (37.4) | 77 (25.8) | 56 (18.6) | 48 (18.2) |  |
| **Overall** | 5382 (58.8) | 2211 (24.8) | 932 (10.7) | 442 (5.7) |  |

Unweighted counts and survey-weighted percentages are reported; row totals reflect unweighted population.
<sup>1</sup> Counts and percentages are pooled across imputations.
<sup>2</sup> Maltreatment types included are physical abuse, sexual abuse, and exposure to domestic violence.
<sup>3</sup> Weighted chi-square test used to calculate p-values.

### Childhood Maltreatment and Mental-Physical Multimorbidity

As shown in Table 3, we found higher odds of all multimorbidity states with each additional childhood maltreatment type reported compared to those reporting no maltreatment in both unadjusted and adjusted analyses. The unadjusted odds ratios (OR) ranged from OR=2.19 (95% CI:1.93-2.48) with 1 reported maltreatment type to OR=9.25 (95% CI:7.6-11.26) with 3 reported maltreatment types for mental-physical multimorbidity. Estimates ranged from OR=1.41 (95% CI:1.35-1.48) to OR=1.99 (95% CI:1.75-2.26) for physical multimorbidity and from OR=1.79 (95% CI:1.58-2.02) to OR=3.94 (95% CI:3.22-4.83) for mental multimorbidity.

**Table 3.**
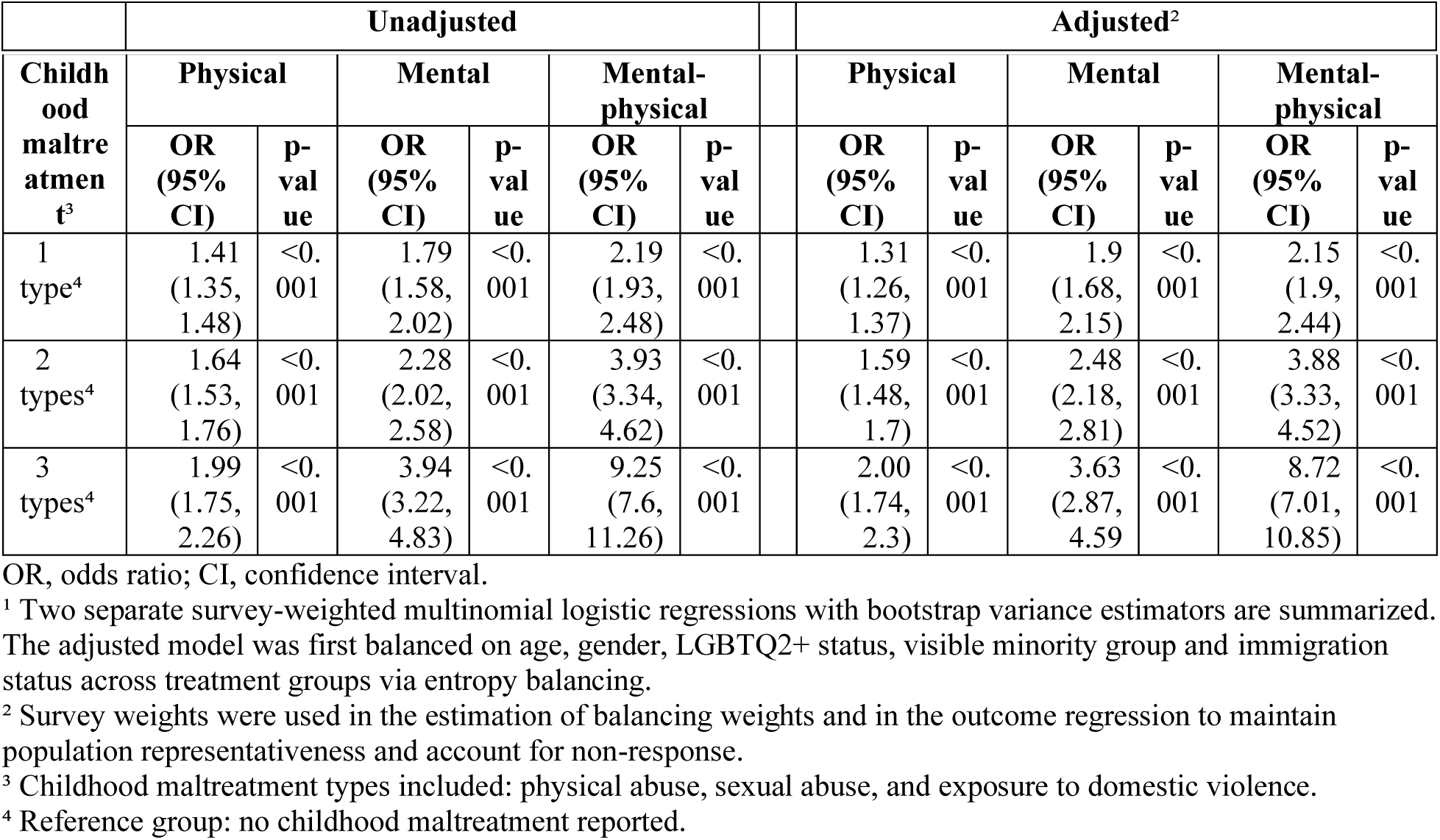
Odds of physical multimorbidity, mental multimorbidity, and mental-physical multimorbidity among Canadian individuals over 20 years of age who reported exposure to 1 or more types of childhood maltreatment compared to those who did not report exposure to any types of childhood maltreatment based on the 2022 Mental Health and Access to Care Survey (MHACS).¹

Associations remained robust after balancing the sample on age, gender, LGBTQ2+ identity, visible minority group and immigration status. Mental-physical multimorbidity exhibited the strongest graded association with adjusted odds ratios (aOR) ranging from aOR=2.15 (95% CI: 1.9-2.44) with 1 maltreatment type to aOR=8.72 (95% CI: 7.01-10.85) with 3 maltreatment types. Odds ratios ranged from aOR=1.31 (95% CI: 1.26-1.37) to aOR=2.00 (95% CI: 1.74-2.3) for physical multimorbidity and from aOR=1.9 (95% CI: 1.68-2.15) to aOR=3.63 (95% CI: 2.87, 4.59) for mental multimorbidity. All associations were statistically significant (p<0.001) with no confidence interval overlap between exposure levels, demonstrating a progressive effect increase across all multimorbidity states with increasing maltreatment exposure (Fig 2).

**Fig 2.**
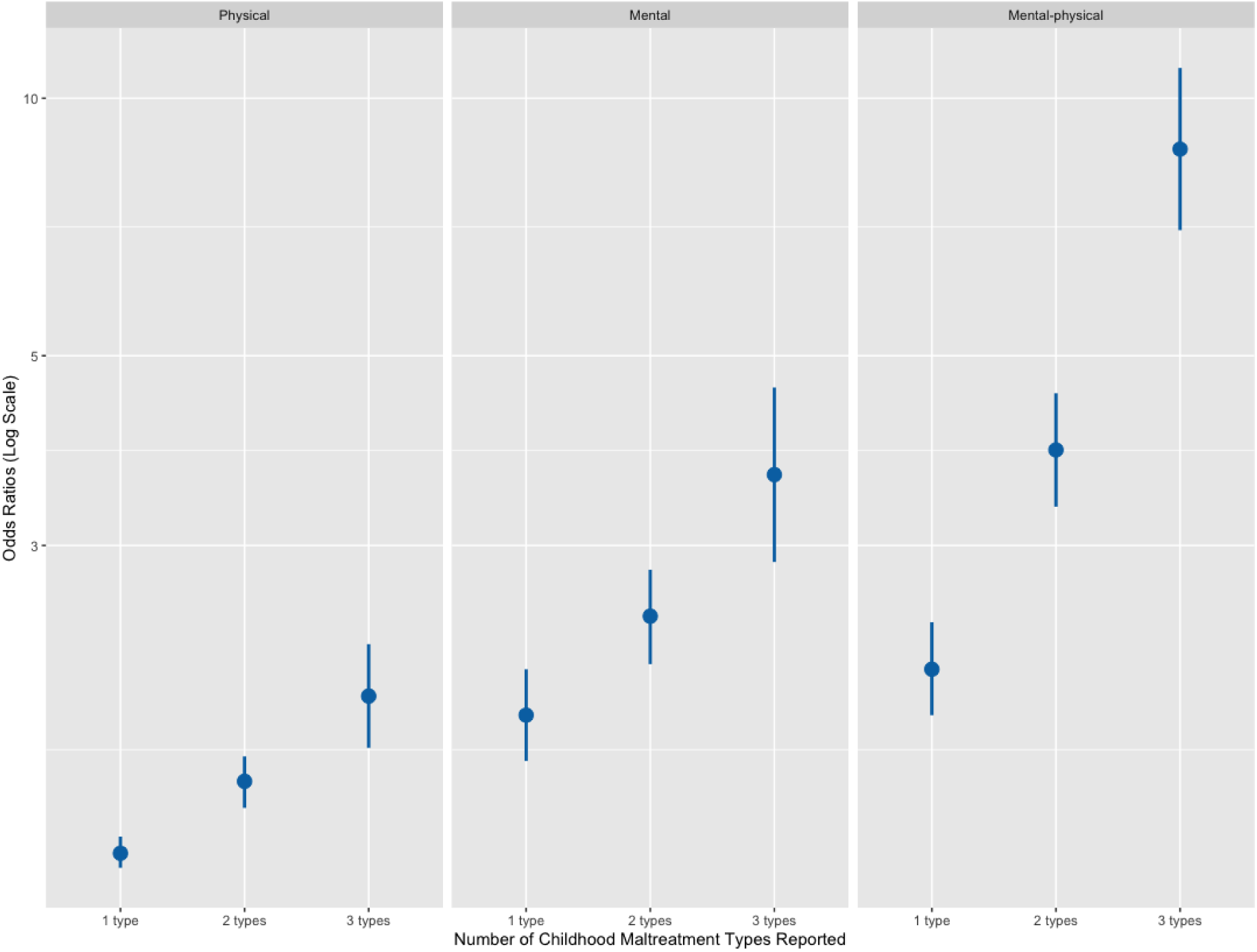
Forest plot of the adjusted relationship (odds ratios) between number of types (physical abuse, sexual abuse, exposure to domestic violence) of childhood maltreatment reported before 16 years of age and multimorbidity among Canadian adults (> 20 years of age) by multimorbidity state based on the 2022 Mental Health and Access to Care Survey (MHACS).

### Effect Modification by Gender

Table 4 presents the gender-specific effect estimates of childhood maltreatment on multimorbidity from a single model with a gender-by-maltreatment interaction. While interactions were not significant at the lowest level of exposure to childhood maltreatment, they were significant for both mental-physical multimorbidity and physical multimorbidity at higher exposure levels. For mental-physical multimorbidity, the effect in women ranged from aOR=2.13 (95% CI:1.81-2.51) with 1 maltreatment type to aOR=6.14 (95% CI:4.90-7.70) with 3 types. In men, associations were comparable at low exposure (aOR=2.21, 95% CI:1.76-2.78) but stronger at high exposure (aOR=13.96, 95% CI:9.58-20.34), indicating magnified gender differences with increasing maltreatment exposure (Fig 3).

**Fig 3.**
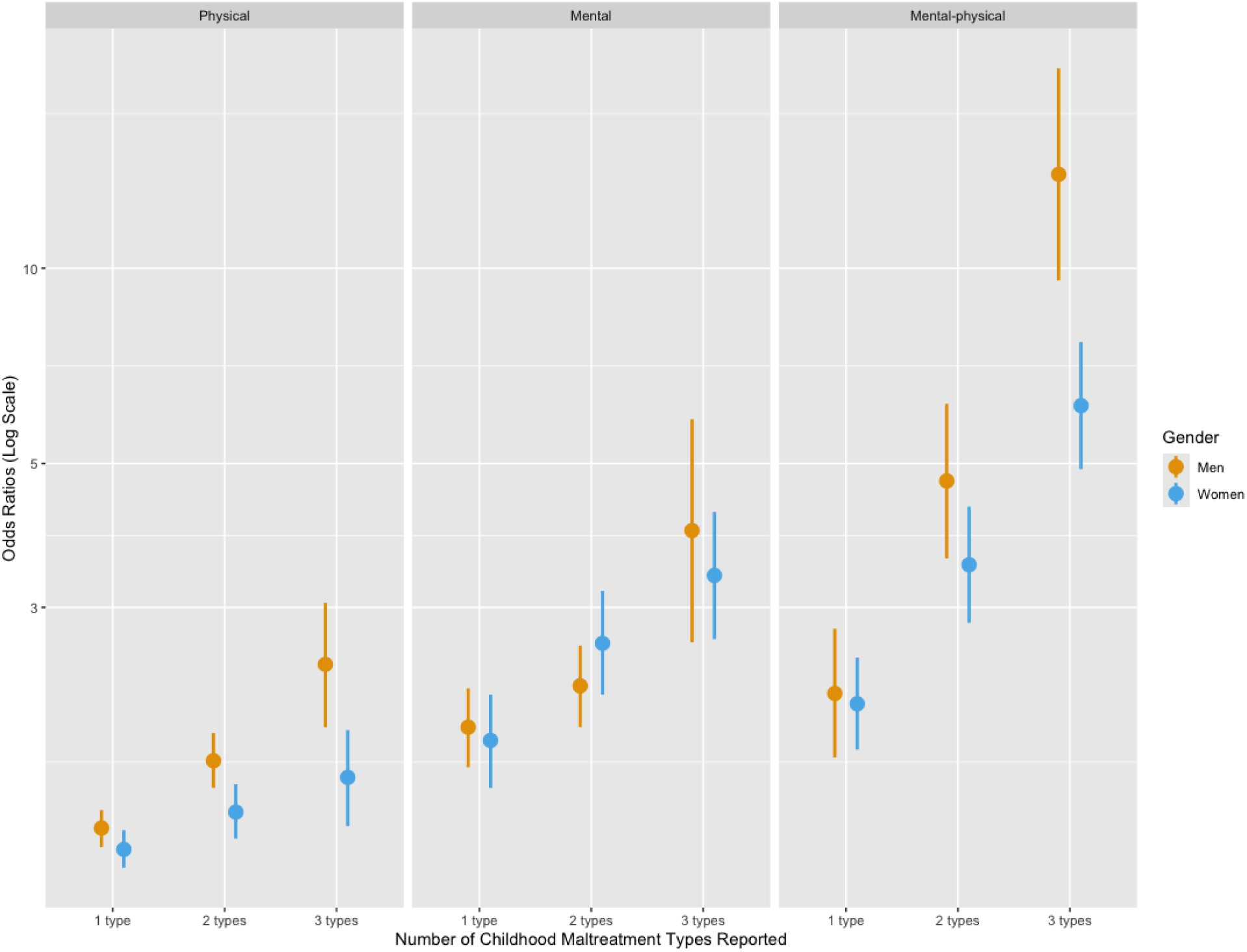
Forest plot of the adjusted relationship (odds ratios) between number of types (physical abuse, sexual abuse, exposure to domestic violence) of childhood maltreatment reported before 16 years of age and multimorbidity among Canadian adults (> 20 years of age) by multimorbidity state and gender based on the 2022 Mental Health and Access to Care Survey (MHACS).

**Table 4.** Gender-specific odds of physical multimorbidity, mental multimorbidity, and mental-physical multimorbidity among Canadian individuals over 20 years of age who reported exposure to 1 or more types of childhood maltreatment compared to those who did not report exposure to any types of childhood maltreatment based on the 2022 Mental Health and Access to Care Survey (MHACS).¹

| Child<br>hood<br>maltr<br>eatme<br>nt<sup>3</sup> | Men |  |  |  |  |  | Women |  |  |  |  |  |
| --- | --- | --- | --- | --- | --- | --- | --- | --- | --- | --- | --- | --- |
|  | Physical |  | Mental |  | Mental-physical |  | Physical |  | Mental |  | Mental-physical |  |
|  | OR<br>(95%<br>CI) | p-<br>val<br>ue | OR<br>(95%<br>CI) | p-<br>val<br>ue | OR (95%<br>CI) | p-<br>val<br>ue | OR<br>(95%<br>CI) | p-<br>val<br>ue | OR<br>(95%<br>CI) | p-<br>val<br>ue | OR<br>(95%<br>CI) | p-<br>val<br>ue |
| 1 type<sup>4</sup> | 1.37<br>(1.28, 1.46) | <0.001 | 1.96<br>(1.7, 2.25) | <0.001 | 2.21<br>(1.76, 2.78) | <0.001 | 1.27<br>(1.19, 1.36) | <0.001 | 1.87<br>(1.58, 2.2) | <0.001 | 2.13<br>(1.81, 2.51) | <0.001 |
| 2 types<sup>4</sup> | 1.74<br>(1.58, 1.92)** | <0.001 | 2.27<br>(1.96, 2.62) | <0.001 | 4.7 (3.57, 6.18) | <0.001 | 1.45<br>(1.32, 1.6)** | <0.001 | 2.64<br>(2.2, 3.18) | <0.001 | 3.49<br>(2.84, 4.29) | <0.001 |
| 3 types<sup>4</sup> | 2.45<br>(1.96, 3.05)** | <0.001 | 3.94<br>(2.65, 5.85) | <0.001 | 13.96<br>(9.58, 20.34)*** | <0.001 | 1.64<br>(1.38, 1.94)** | <0.001 | 3.36<br>(2.68, 4.21) | <0.001 | 6.14<br>(4.9, 7.7)*** | <0.001 |

OR, odds ratio; CI, confidence interval; \*, significant interaction at 0.05 level; \*\*, significant interaction at 0.01 level; \*\*\*, significant interaction at 0.001 level.
<sup>1</sup> Results from a single survey-weighted multinomial logistic regression with a bootstrap variance estimator are summarized. The model was first balanced on age, gender, LGBTQ2+ status, visible minority group and immigration status across treatment groups via entropy balancing; the outcome regression included an interaction between gender and childhood maltreatment.
<sup>2</sup> Survey weights were used in the estimation of balancing weights and in the outcome regression to maintain population representativeness and account for non-response.
<sup>3</sup> Childhood maltreatment types included: physical abuse, sexual abuse, and exposure to domestic violence.
<sup>4</sup> Reference group: no childhood maltreatment reported.

Associations were more modest for physical multimorbidity. Effect estimates in women ranged from aOR=1.27 (95% CI:1.19-1.36) with 1 reported maltreatment type to aOR=1.64 (95% CI:1.38-1.94) with 3 types. In men, effect estimates ranged from aOR=1.37 (95% CI:1.28-1.46) with 1 maltreatment type to aOR=2.45 (95% CI:1.96-3.05) with 3. Effect modification was weaker and non-significant for mental multimorbidity across all exposure levels ranging from aOR=1.87 (95% CI:1.58-2.2) to aOR=3.36 (95% CI:2.68-4.21) in women and from aOR=1.96 (95% CI:1.7-2.25) to aOR=3.94 (95% CI:2.65-5.85) in men.

### Cluster-Specific Analyses

In cluster-specific models, findings were consistent with the main analysis (Table 5 and Fig 4). However, patterns varied significantly when effect modification was considered (Table 6). In the mental-cardiometabolic cluster, odds of mental-physical multimorbidity were much higher in men (aOR=7.28, 95% CI:2.81-18.87) than women (aOR=0.62, 95% CI:0.29-1.33) but converged at higher levels of exposure (Fig 5). In the mental-inflammatory cluster, effect modification was not significant except in physical multimorbidity where women exhibited higher odds of physical multimorbidity with 3 maltreatment types (aOR=2.44, 95% CI:1.87-3.18 in women; aOR=1.39, 95% CI:0.91-2.13 in men). In the mental-somatic cluster, stronger associations were observed in women with 1 maltreatment type (aOR =2.82, 95% CI:2.14-3.71 in women; aOR=0.99, 95% CI:0.65-1.5 in men). However, associations were stronger in men with 3 maltreatment types (aOR=6.45, 95% CI:4.57-9.1 in women; aOR=14.61, 95% CI:9.92-21.52 in men).

**Fig 4.**
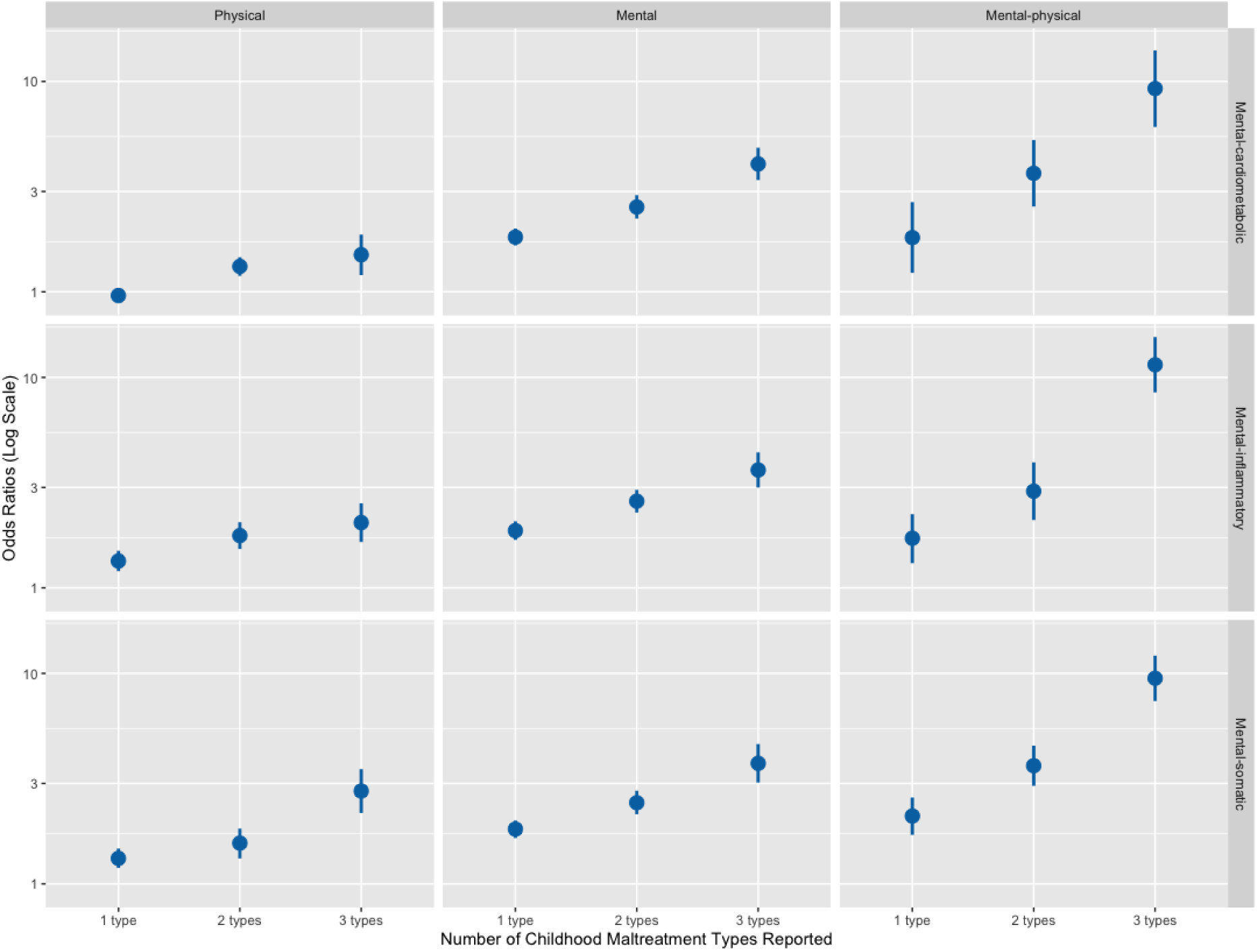
Forest plot of the adjusted relationship (odds ratios) between number of types (physical abuse, sexual abuse, exposure to domestic violence) of childhood maltreatment reported before 16 years of age and multimorbidity among Canadian adults (> 20 years of age) by multimorbidity state and mental-physical disease cluster based on the 2022 Mental Health and Access to Care Survey (MHACS).

**Fig 5.**
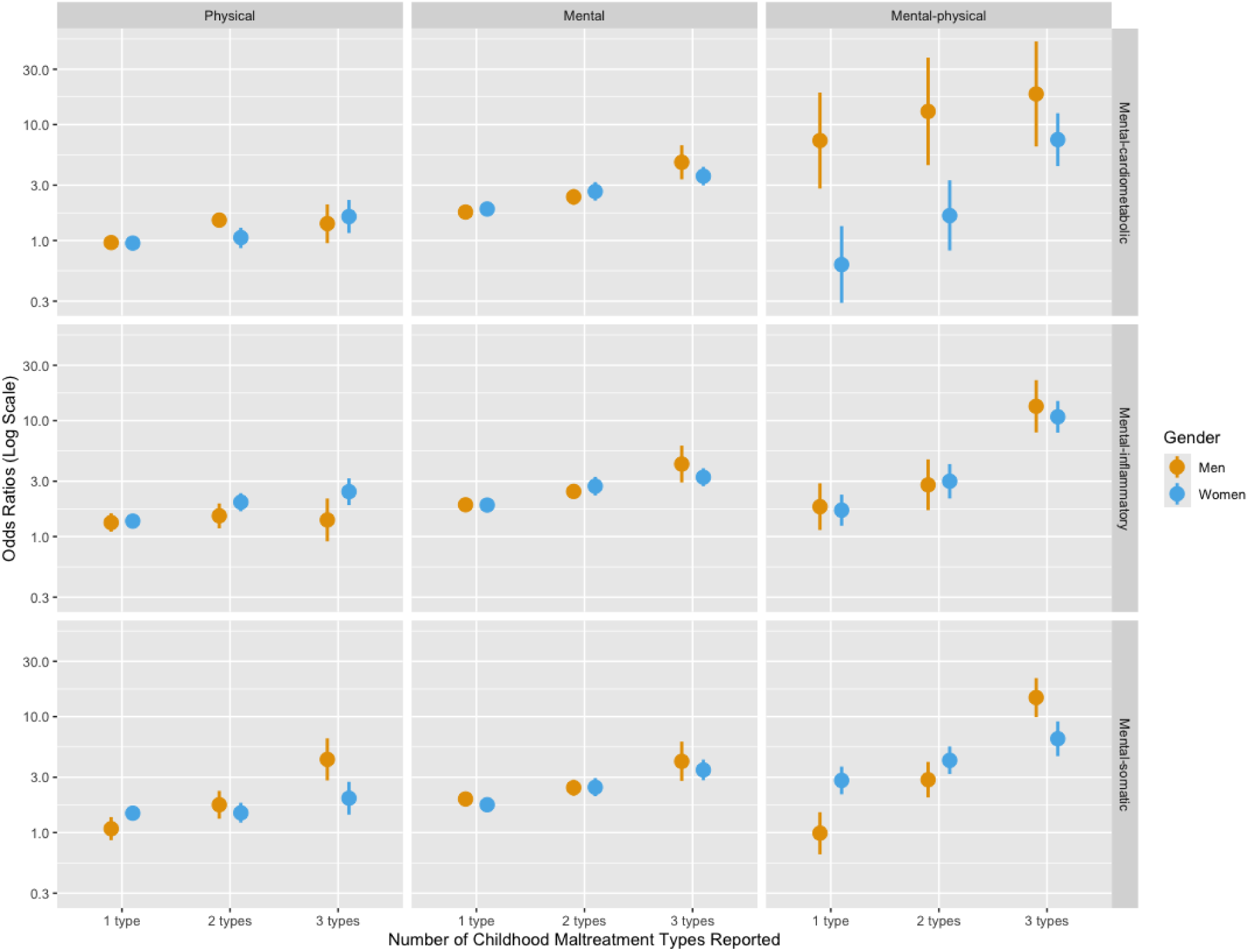
Forest plot of the adjusted relationship (odds ratios) between number of types (physical abuse, sexual abuse, exposure to domestic violence) of childhood maltreatment reported before 16 years of age and multimorbidity among Canadian adults (> 20 years of age) by multimorbidity state, mental-physical disease cluster, and gender based on the 2022 Mental Health and Access to Care Survey (MHACS).

**Table 5.** Odds of cluster-specific physical multimorbidity, mental multimorbidity, and mental-physical multimorbidity among Canadian individuals over 20 years of age who reported exposure to 1 or more types of childhood maltreatment compared to those who did not report exposure to any types of childhood maltreatment based on the 2022 Mental Health and Access to Care Survey (MHACS).¹

|  | Unadjusted |  |  |  |  |  | Adjusted<sup>2</sup> |  |  |  |  |  |
| --- | --- | --- | --- | --- | --- | --- | --- | --- | --- | --- | --- | --- |
| Mental-cardiometabolic multimorbidity<sup>2</sup> |  |  |  |  |  |  |  |  |  |  |  |  |
| Child<br>hood<br>maltre<br>atmen<br>t<sup>3</sup> | Physical |  | Mental |  | Mental-physical |  | Physical |  | Mental |  | Mental-physical |  |
|  | OR<br>(95%<br>CI) | p-<br>val<br>ue | OR<br>(95%<br>CI) | p-<br>val<br>ue | OR<br>(95%<br>CI) | p-<br>val<br>ue | OR<br>(95%<br>CI) | p-<br>val<br>ue | OR<br>(95%<br>CI) | p-<br>val<br>ue | OR<br>(95%<br>CI) | p-<br>val<br>ue |
| 1<br>type<sup>4</sup> | 1.05<br>(0.98,<br>1.13) | 0.17 | 1.71<br>(1.56,<br>1.88) | <0.001 | 2.2<br>(1.48,<br>3.26) | <0.001 | 0.96<br>(0.89,<br>1.03) | 0.27 | 1.82<br>(1.66,<br>2.00) | <0.001 | 1.81<br>(1.23,<br>2.67) | 0.004 |
| 2<br>types<sup>4</sup> | 1.43<br>(1.29,<br>1.6) | <0.001 | 2.4<br>(2.12,<br>2.72) | <0.001 | 3.85<br>(2.66,<br>5.58) | <0.001 | 1.32<br>(1.19,<br>1.46) | <0.001 | 2.53<br>(2.23,<br>2.88) | <0.001 | 3.66<br>(2.54,<br>5.27) | <0.001 |
| 3<br>types<sup>4</sup> | 1.26<br>(1.01,<br>1.56) | 0.004 | 4.21<br>(3.63,<br>4.89) | <0.001 | 11.63<br>(7.83,<br>17.29) | <0.001 | 1.5<br>(1.2,<br>1.87) | 0.002 | 4.05<br>(3.4,<br>4.84) | <0.001 | 9.25<br>(6.08,<br>14.06) | <0.001 |
| Mental-inflammatory multimorbidity<sup>2</sup> |  |  |  |  |  |  |  |  |  |  |  |  |
| Child<br>hood<br>maltre<br>atmen<br>t<sup>3</sup> | Physical |  | Mental |  | Mental-physical |  | Physical |  | Mental |  | Mental-physical |  |
|  | OR<br>(95%<br>CI) | p-<br>val<br>ue | OR<br>(95%<br>CI) | p-<br>val<br>ue | OR<br>(95%<br>CI) | p-<br>val<br>ue | OR<br>(95%<br>CI) | p-<br>val<br>ue | OR<br>(95%<br>CI) | p-<br>val<br>ue | OR<br>(95%<br>CI) | p-<br>val<br>ue |

| 1 type<sup>4</sup> | 1.39<br>(1.24, 1.56) | <0.001 | 1.75<br>(1.59, 1.93) | <0.001 | 1.76<br>(1.32, 2.34) | <0.001 | 1.34<br>(1.2, 1.5) | <0.001 | 1.87<br>(1.69, 2.07) | <0.001 | 1.72<br>(1.31, 2.24) | <0.001 |
| --- | --- | --- | --- | --- | --- | --- | --- | --- | --- | --- | --- | --- |
| 2 types<sup>4</sup> | 1.67<br>(1.45, 1.93) | <0.001 | 2.41<br>(2.14, 2.71) | <0.001 | 2.93<br>(2.1, 4.09) | <0.001 | 1.77<br>(1.53, 2.05) | <0.001 | 2.58<br>(2.28, 2.92) | <0.001 | 2.88<br>(2.1, 3.95) | <0.001 |
| 3 types<sup>4</sup> | 2.29<br>(1.87, 2.79) | <0.001 | 3.85<br>(3.29, 4.5) | <0.001 | 13.86<br>(10.61, 18.09) | <0.001 | 2.04<br>(1.65, 2.52) | <0.001 | 3.63 (3, 4.4) | <0.001 | 11.49<br>(8.49, 15.55) | <0.001 |
| **Mental-somatic multimorbidity<sup>2</sup>** |  |  |  |  |  |  |  |  |  |  |  |  |
| Child hood maltreatment<sup>3</sup> | Physical |  | Mental |  | Mental-physical |  | Physical |  | Mental |  | Mental-physical |  |
|  | OR (95% CI) | p-value | OR (95% CI) | p-value | OR (95% CI) | p-value | OR (95% CI) | p-value | OR (95% CI) | p-value | OR (95% CI) | p-value |
| 1 type<sup>4</sup> | 1.33<br>(1.19, 1.49) | <0.001 | 1.74<br>(1.58, 1.91) | <0.001 | 1.8<br>(1.47, 2.2) | <0.001 | 1.32<br>(1.19, 1.47) | <0.001 | 1.82<br>(1.65, 2.0) | <0.001 | 2.1<br>(1.71, 2.57) | <0.001 |
| 2 types<sup>4</sup> | 1.55<br>(1.31, 1.85) | <0.001 | 2.3<br>(2.03, 2.61) | <0.001 | 3.42<br>(2.7, 4.32) | <0.001 | 1.56<br>(1.32, 1.83) | <0.001 | 2.43<br>(2.14, 2.77) | <0.001 | 3.64<br>(2.92, 4.54) | <0.001 |
| 3 types<sup>4</sup> | 2.95<br>(2.32, 3.74) | <0.001 | 4.17<br>(3.49, 4.98) | <0.001 | 8.95<br>(7.02, 11.42) | <0.001 | 2.76<br>(2.17, 3.51) | 0.002 | 3.74<br>(3.03, 4.62) | <0.001 | 9.48<br>(7.39, 12.16) | <0.001 |

OR, odds ratio; CI, confidence interval.
<sup>1</sup> Separate survey-weighted multinomial logistic regressions using a bootstrap variance estimator are summarized. The adjusted models were first balanced on age, gender, LGBTQ2+ status, visible minority group and immigration status across treatment groups via entropy balancing. Survey weights were used in the estimation of balancing weights and in the outcome regression to maintain population representativeness and account for non-response.
<sup>2</sup> Diseases included in each cluster are mental-inflammatory: asthma, chronic obstructive pulmonary disease, arthritis, inflammatory bowel disease; mental-somatic: chronic fatigue syndrome, back pain, chronic migraines, fibromyalgia; mental-cardiometabolic: diabetes, heart disease, and hypertension. All mental health conditions were included in each cluster.
<sup>3</sup> Childhood maltreatment types included: physical abuse, sexual abuse, and exposure to domestic violence.
<sup>4</sup> Reference group: no childhood maltreatment reported.

**Table 6.**
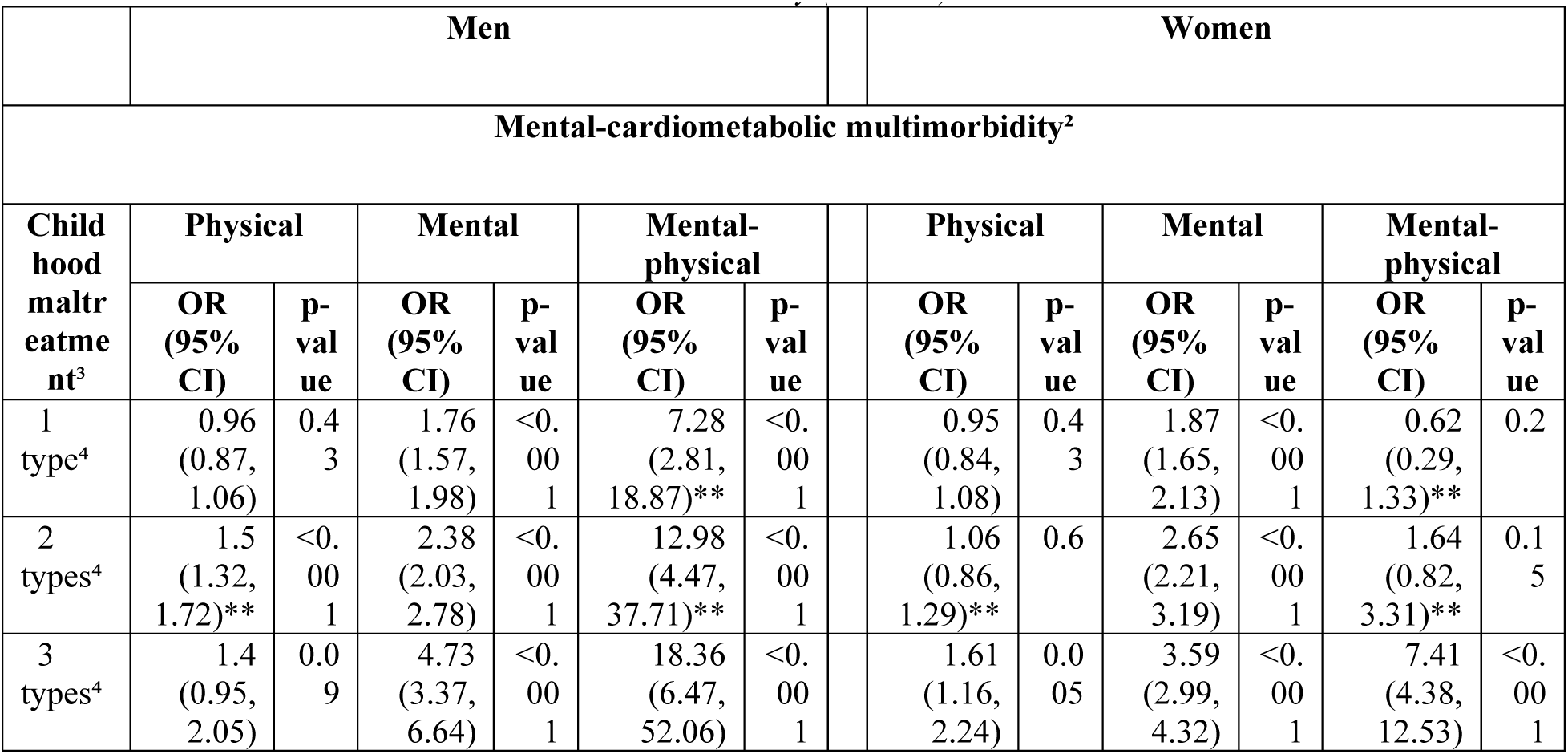
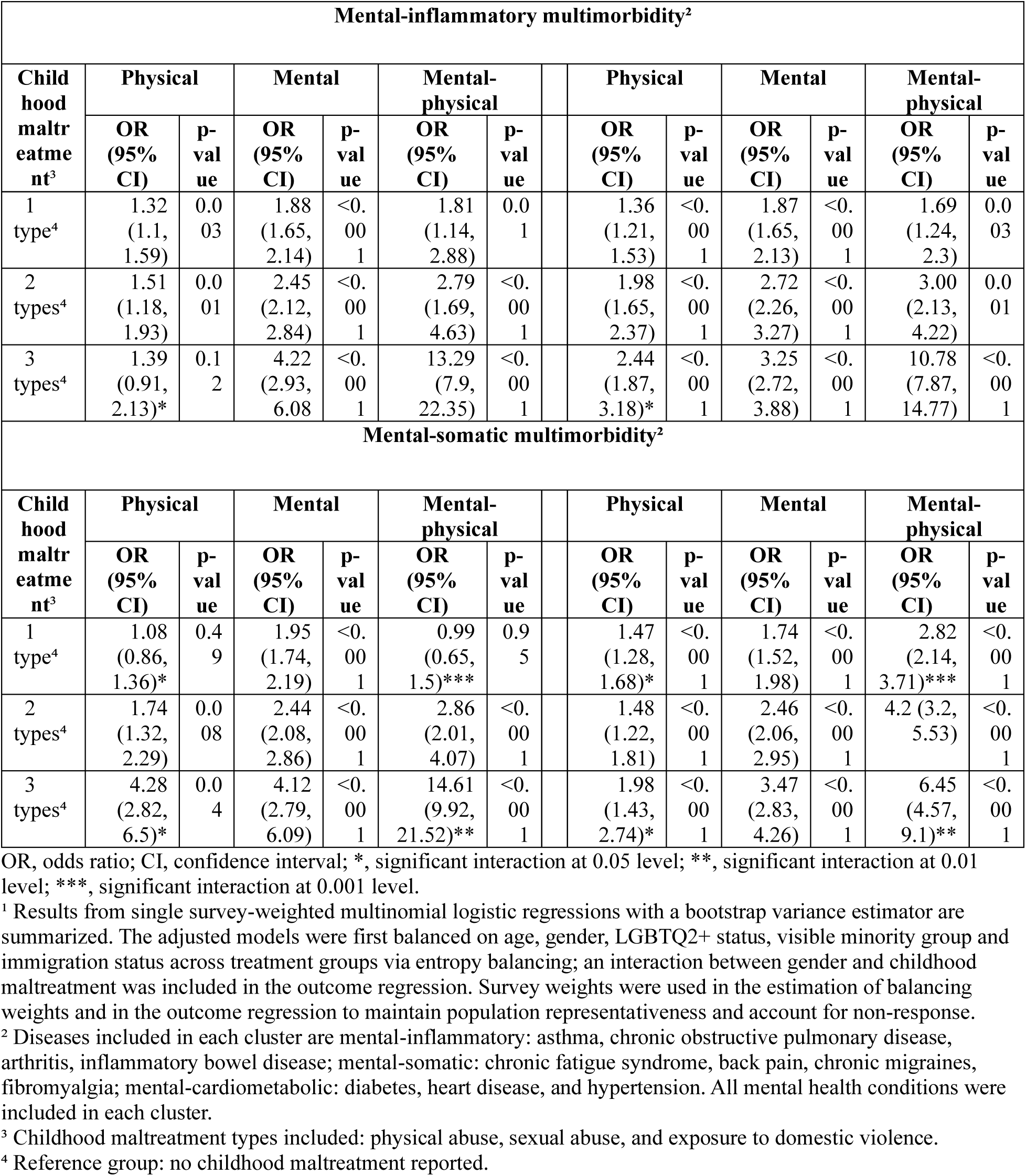
Gender-specific odds of cluster-specific physical multimorbidity, mental multimorbidity, and mental- physical multimorbidity among Canadian individuals over 20 years of age who reported exposure to 1 or more types of childhood maltreatment compared to those who did not report exposure to any types of childhood maltreatment based on the 2022 Mental Health and Access to Care Survey (MHACS).¹

### Subtype-Specific Analyses

Maltreatment was associated with higher unadjusted and adjusted odds of all multimorbidity states across the subtype-specific models, but associations with mental-physical multimorbidity were strongest (Table 7). The association between sexual abuse and mental-physical multimorbidity was stronger in men (aOR=3.07, 95% CI:2.50-3.77) than women (aOR=1.66, 95% CI:1.41-1.95), whereas the association with physical abuse was stronger in women (aOR=2.95, 95% CI:2.59-3.36; aOR=2.07, 95% CI:1.74-2.45 in men) (Table 8). Exposure to domestic violence was associated with increased odds of mental-physical multimorbidity in men (aOR=2.17, 95% CI:1.76-2.66) but decreased odds in women (aOR=0.79, 95% CI:0.66-0.95).

**Table 7.**
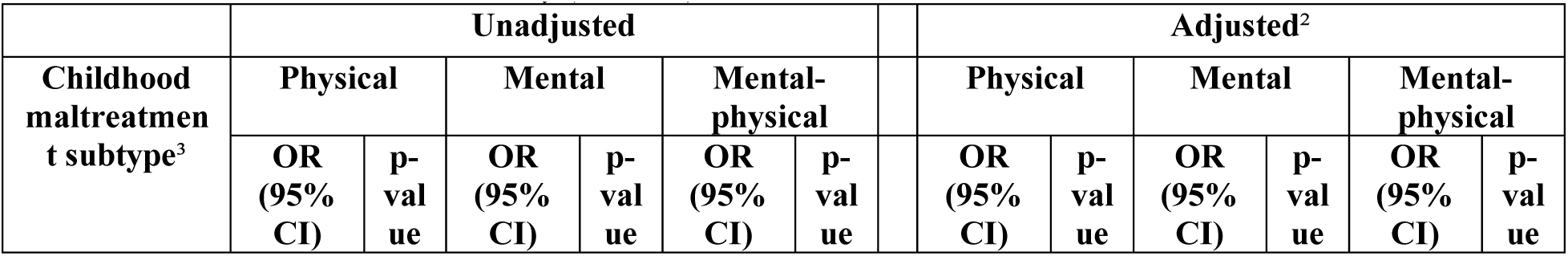
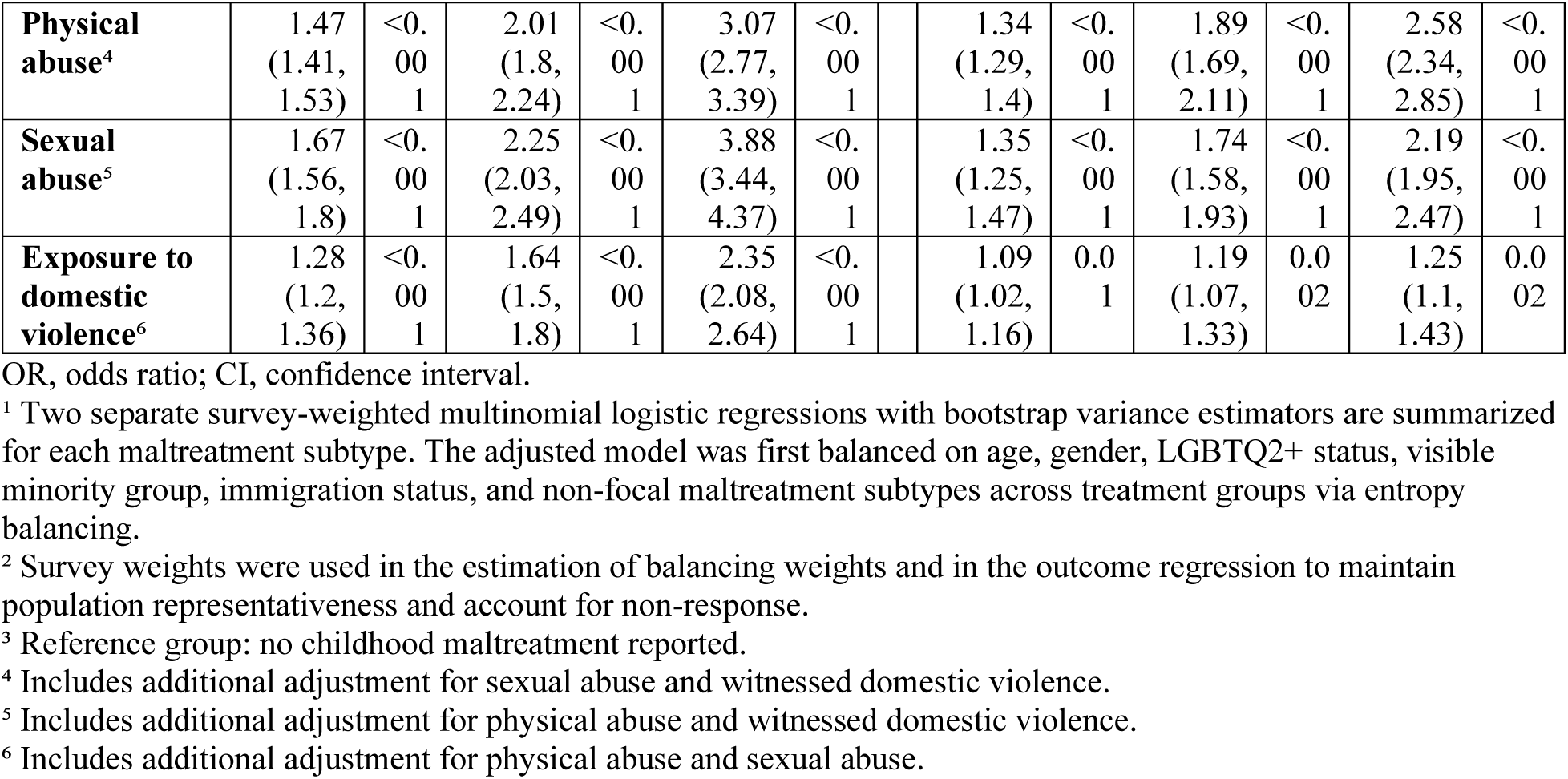
Odds of physical multimorbidity, mental multimorbidity, and mental-physical multimorbidity among Canadian individuals over 20 years of age who reported exposure to physical abuse, sexual abuse, or exposure to domestic violence compared to those who did not report subtype-specific childhood maltreatment based on the 2022 Mental Health and Access to Care Survey (MHACS).¹

**Table 8.**
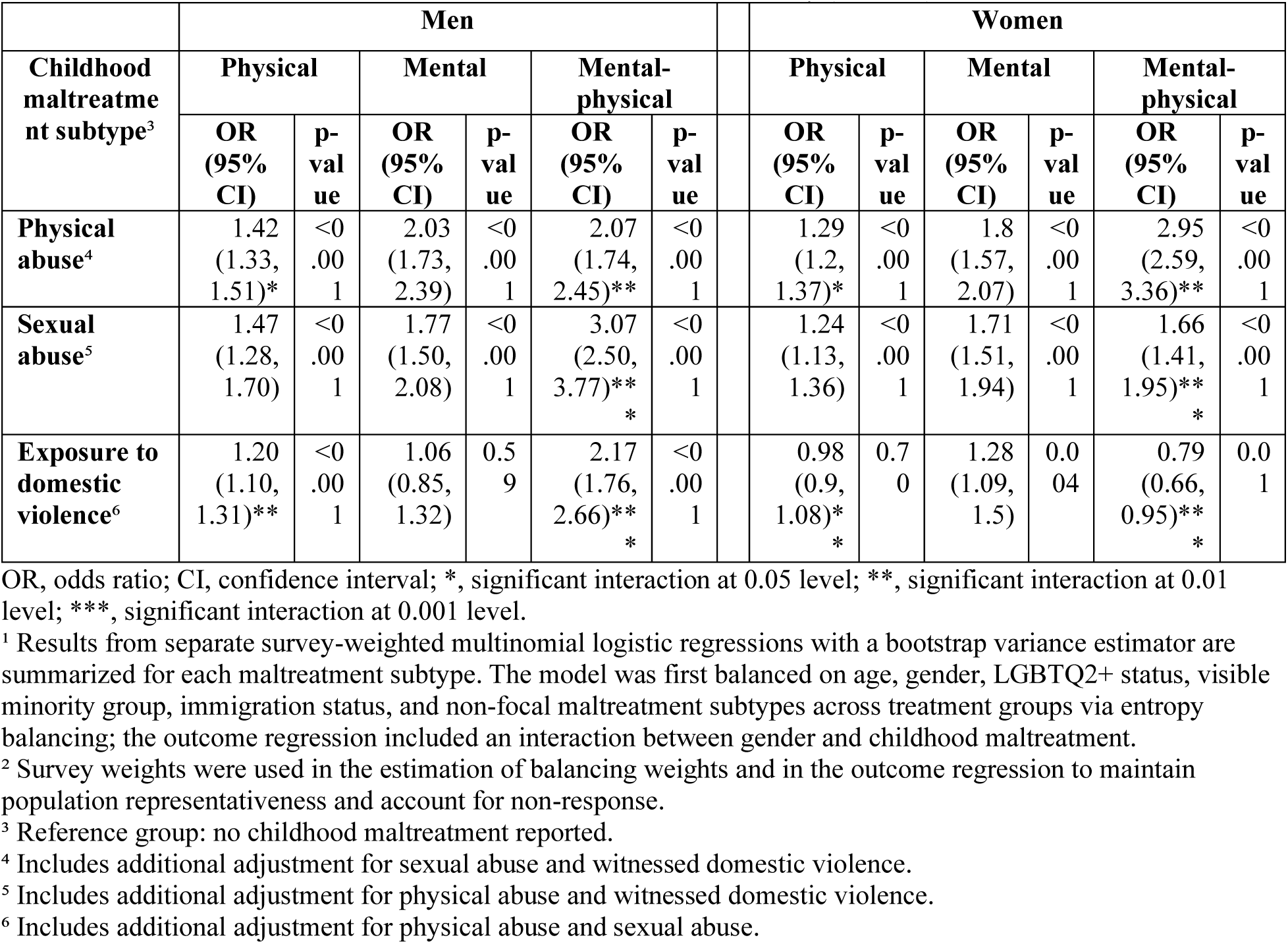
Gender-specific odds of physical multimorbidity, mental multimorbidity, and mental-physical multimorbidity among Canadian individuals over 20 years of age who reported exposure to physical abuse, sexual abuse, or exposure to domestic violence compared to those who did not report subtype-specific childhood maltreatment based on the 2022 Mental Health and Access to Care Survey (MHACS).¹

## DISCUSSION

Our findings demonstrate a robust dose-response association between childhood maltreatment and adulthood multimorbidity with particularly strong effects for mental-physical multimorbidity. Although effect sizes varied, this pattern persisted in gender-specific analyses and held across mental-cardiometabolic, mental-inflammatory, and mental-somatic clusters.

These findings are consistent with prior Canadian research. Using 2012 CCHS data and an identically structured exposure variable, England-Mason et al. (2018) reported a similar dose-response pattern. For co-occurring mental and physical conditions (excluding chronic pain/fatigue), their adjusted odds ratios ranged from 1.98 (95% CI:1.38-2.85) with 1 maltreatment type to 6.51 (95% CI:3.09-13.70) with all types. For co-occurring mental and chronic pain/fatigue conditions, they ranged from 1.44 (95% CI: 1.21-1.73) to 3.38 (95% CI:1.94-5.87), and for the co-occurrence of all three conditions they ranged from 2.63 (95% CI: 1.93-3.58) to 10.96 (95% CI:6.12-19.64). Comparatively, our adjusted estimates range from 2.15 (95% CI:1.90-2.44) with 1 type to 8.72 (95% CI:7.01-10.85) with all types for mental-physical multimorbidity, and from 2.1 (95% CI:1.71-2.57) to 9.48 (95% CI:7.39-12.16) for mental-somatic multimorbidity. Although not directly comparable due to England-Mason et al.’s distinct outcome specification and additional behavioural and socioeconomic adjustments, the overall dose-response pattern is consistent. More broadly, our results align with related research in Canada and the UK demonstrating strong associations between childhood maltreatment and both mental-cardiometabolic (Souama et al., 2023) and mental-inflammatory multimorbidity (Wan et al., 2022; O’Mahony et al., 2024) as well as existing biopsychosocial evidence linking childhood maltreatment to adulthood morbidity (Hughes et al., 2017; Metzler et al., 2016; Baltramonaityte et al., 2023).

We also identified novel gender-specific patterns with men showing particularly strong associations at higher maltreatment exposure levels (127.4% stronger in men reporting all maltreatment types). Importantly, these gender differences varied across disease clusters and subtype-specific exposures. Effect modification was strongest in the mental-cardiometabolic cluster (147.8% stronger in men reporting all maltreatment types), weaker in mental-somatic (126.5% stronger in men reporting all maltreatment types), and minimal in the mental- inflammatory cluster (23.2% stronger in men reporting all maltreatment types). Subtype-specific exposure models also showed a stronger association between sexual abuse and mental-physical multimorbidity in men (184.9% stronger in men), whereas physical abuse was more strongly associated with mental-physical multimorbidity in women (142.5% stronger in women).

Although there is a paucity of comparable research in Canada, our findings contrast with a recent meta-analysis in the UK which did not find any evidence of sex-based differences in the association between childhood maltreatment and co-occurring depression and cardiometabolic disease (Souama et al., 2023). While the biological mechanisms involved in mental-physical multimorbidity may differ from those driving the development of individual conditions, it is noteworthy that these findings also contrast with prior Canadian research which has reported stronger associations between maltreatment and individual mental (Gallo et al., 2018) and physical (Wan et al., 2022) morbidities in women than in men. Direct comparisons should be made cautiously given that these studies adjusted for additional socioeconomic and behavioural covariates omitted from our analytic model. Our results are, however, generally consistent with recent evidence from Ontario and British Columbia indicating that the incidence and prevalence of multimorbidity are increasing at greater rates among Canadian men (Kone et al., 2021; Ferris et al., 2025).

While gender-specific pathways linking childhood maltreatment to mental-physical multimorbidity remain underexplored in Canada, several potential mechanisms may explain the gender differences observed in our analysis. Recent Canadian estimates based on MHACS indicate higher prevalence of mood and anxiety disorders among women and higher prevalence of substance use disorders among men (Stephenson, 2023). Meanwhile, established evidence in the US has linked increased incidence of internalizing disorders (e.g. anxiety and mood disorders) in women to inflammatory and chronic pain conditions, and increased incidence of externalizing disorders (e.g. substance use disorders) in men to cardiometabolic diseases (Needham & Hill, 2010). Alongside evidence indicating that men are more likely to exhibit impaired cardiometabolic functioning while women are more likely to exhibit neuroendocrine and immune dysregulation (Longpré-Poirer et al., 2022), these associations provide some plausible explanations for our findings. Other research suggests that men may be more likely to engage in health-harming behaviours associated with multimorbidity (Punjani et al., 2018) and less likely to access healthcare services compared to women (Thompson et al., 2016). Childhood maltreatment may exacerbate these existing biological and behavioural vulnerabilities in men, resulting in an accumulation of risk factors for multimorbidity across the life course. These mechanisms alone, however, are likely insufficient in explaining the increased effect sizes seen among men in our analysis. Further research is needed to understand the gender-specific effects of childhood maltreatment on mental-physical multimorbidity with attention to specific maltreatment subtypes.

## Strengths and Limitations

To our knowledge, this study is the first in Canada to report gender-specific associations between childhood maltreatment and mental-physical multimorbidity using a nationally representative sample. However, several limitations must be considered. First, the cross-sectional design prohibited temporal investigation of multimorbidity onset and precludes causal interpretation.

While our findings suggest that childhood maltreatment is associated with increased odds of mental-physical multimorbidity, they do not suggest that this relationship is causal. Second, relevant early life covariates such as childhood household conditions were not measured and no proxy measures could be identified. Additionally, the broad analytic focus of our study prohibited disease-specific adjustments. Although post-hoc E-value estimation indicates that unmeasured confounders would need to both be more prevalent among those exposed to maltreatment and increase the risk of mental-physical multimorbidity by a factor of 2.29 for those reporting 1 type of maltreatment and 16.92 for those reporting all types to fully explain the observed associations, some residual confounding is anticipated. Third, it is important to note that the gender variable in the MHACS dataset includes non-binary individuals who were randomly imputed into the “Men+” and “Women+” categories and could not be re-identified (Statistics Canada, 2023), and that men were underrepresented at the highest exposure level.

Although we adjusted for LGBTQ2+ identity and the latter is reflected in the uncertainty associated with our estimates, these factors should be considered alongside possible misclassification bias when interpreting our gender-specific findings.

There are also limitations related to our exposure and outcome definitions. Our main exposure variable aligns with existing maltreatment surveillance frameworks in Canada and implicitly captures aspects of both cumulative and subtype-specific exposure, but it does not include neglect or emotional maltreatment due to data limitations. While sub-analyses were performed as a preliminary effort to disentangle subtype-specific contributions, complex interactions were not explored. Finally, it was not possible to include or investigate all potentially relevant conditions associated with early life trauma. Conditions with a significant autoimmune component (e.g., arthritis, IBD), for example, were grouped with inflammatory diseases while eating disorders, post-traumatic stress disorder, and personality disorders represent notable omissions from the included mental health conditions.

## Conclusion

While our findings indicate that childhood maltreatment is associated with increased odds of mental-physical multimorbidity, they do not suggest that early adversity leads deterministically to multimorbidity. To support intervention development, longitudinal studies should aim to clarify the gender-specific biopsychosocial pathways linking childhood maltreatment to mental-physical multimorbidity using linked administrative data and identify opportunities for prevention and mitigation. This work should investigate possible mediating factors such as coping strategies, health-related behaviours and socioeconomic achievement. To prevent the progression of morbidity into multimorbidity and multimorbidity into mental-physical multimorbidity, more work is needed to understand the related onset patterns. By preventing further harm and creating additional supports for those affected by childhood maltreatment, it may be possible to reduce the public health burden of mental-physical multimorbidity.

## Data Availability

All data used in this analysis are publicly available and were obtained through the Abacus Data Network.

https://abacus.library.ubc.ca/dataset.xhtml?persistentId=hdl:11272.1/AB2/YIBA26

